# Monoclonal antibody and antiviral therapy for treatment of mild-to-moderate COVID-19 in pediatric patients

**DOI:** 10.1101/2022.03.16.22272511

**Authors:** Surabhi B. Vora, Janet A. Englund, Indi Trehan, Alpana Waghmare, Ada Kong, Amanda Adler, Danielle M. Zerr

## Abstract

The recent surge of SARS-CoV-2 Omicron variant (B.1.1.529) coincided with new treatment options for mild-to-moderate Covid-19 in high-risk adolescents and adults. In this report we describe patient characteristics, treatment-related process measures and outcomes associated with early Covid-19 therapy in high-risk pediatric patients.

## Introduction

The recent surge of SARS-CoV-2 Omicron variant (B.1.1.529) coincided with new treatment options for mild-to-moderate Covid-19 in high-risk adolescents and adults. Though the oral antiviral nirmatrelvir/ritonavir and the monoclonal antibody sotrovimab [1] were approved under Emergency Use Authorization (EUA) for children over 12 years of age, the studies leading up to this approval did not include any pediatric patients [2, 3]. A recent publication demonstrated a three-day course of remdesivir reduces hospitalization rates in high-risk outpatients including younger children [4], but data around the use of any of these agents in children is extremely limited.

Based on Food and Drug Administration (FDA) EUA, pediatric-specific guidance [5], and national prioritization schema [6], our institution has prioritized monoclonal antibodies and early antivirals for severely immunocompromised and incompletely vaccinated patients with additional risk factors including severe obesity, medical complexity with respiratory technology dependence, or multiple other risk factors. Almost all SARS-CoV-2 strains were Omicron variant in our region by late December, 2021 [7]. In this report we describe patient characteristics, treatment-related process measures and outcomes associated with early Covid-19 therapy in high-risk pediatric patients.

## Methods

Patients for whom Covid-19 treatment was requested between 12/22/2021-1/30/2022 were reviewed by our hospital’s Covid-19 Therapeutics Committee with Pediatric Infectious Diseases, Emergency Medicine, and Pharmacy expertise. To determine which treatment to offer we considered each patient’s age, location, ability to tolerate oral treatment, potential drug interactions, underlying renal and liver function and current infusion center or inpatient bed availability. Following IRB approval, medical records were reviewed to assess patient demographics, underlying conditions, SARS-CoV-2 vaccination status, days since first symptom or positive test (PCR or rapid antigen), medication adherence, treatment associated adverse events, and Emergency Department (ED) visits and hospitalizations within 7 days after request intake. Assessment of baseline renal and hepatic function was recommended prior to remdesivir administration.

## Results

Requests for early treatment for 94 patients were reviewed; 66 (70%) received approval for sotrovimab, nirmatrelvir/ritonavir, or remdesivir (Table 1). Immunocompromised patients comprised most requests (66%), with malignancy being the most frequent immunocompromising condition. The most common reasons for denial of therapy were fully vaccinated status and not being considered in the highest risk categories. Self-identified race and ethnicity categorizations were generally similar between those for whom therapy was requested versus approved. Therapy was not given despite approval to 19 patients, most often due to family refusal and improving symptoms (Table 2). Remdesivir treatment occurred most frequently in the inpatient setting while sotrovimab was administered more often in the outpatient infusion center. Adverse events were rare following all agents; no patients had a rise in alanine aminotransferase or creatinine after remdesivir. There were three new (not treatment related) hospitalizations in the follow-up period for potentially Covid-19 related disease, including one who did was not approved for treatment, one who was approved for sotrovimab but did not receive it, and one after treatment with remdesivir (see Table 2 for details). Two additional patients had ED visits, both after therapy (1 sotrovimab, 1 nirmatrelvir/ritonavir); one patient each with shortness of breath and chest pain. Both were discharged in good condition.

**Table 1.** Characteristics of patients for whom COVID-19 therapy was requested and approved.

| **Patient Characteristics** | **Therapy Requested (N, %)**<br>**N=94** | **Therapy Approved (N, %)**<br>**N=66** |
| --- | --- | --- |
| **Median age in years (range)** | 13 (0.6-24) | 12 (0.6-24) |
| <5 years | 16 (17) | 12 (18) |
| 5-11 years | 20 (21) | 13 (20) |
| 12-17 years | 49 (52) | 33 (50) |
| 18+ years | 9 (10) | 8 (12) |
| **Gender** |  |  |
| Female | 43 (46) | 30 (45) |
| **Ethnicity** |  |  |
| Hispanic | 33 (35) | 24 (36) |
| **Race** |  |  |
| White or Caucasian | 41 (44) | 27 (41) |
| Black or African-American | 5 (5) | 4 (6) |
| Asian | 5 (5) | 2 (3) |
| Hawaiian/Pacific-Islander | 4 (4) | 4 (6) |
| Alaska Native/American-Indian | 3 (3) | 3 (5) |
| Multiple | 7 (7) | 5 (7) |
| Other* | 23 (24) | 19 (29) |
| Unknown/Refused | 6 (6) | 2 (3) |
| **Underlying condition(s)\*\*** |  |  |
| Immunocompromised | 62 (66) | 45 (68) |
| Malignancy | 29 (31) | 23 (35) |
| Solid organ transplant | 9 (10) | 7 (11) |
| Hematopoietic cell transplant | 4 (4) | 3 (5) |
| Primary immunodeficiency | 6 (6) | 5 (8) |
| Rheumatologic condition | 5 (5) | 3 (5) |
| Other immunocompromise | 9 (10) | 4 (6) |
| Obesity | 13 (14) | 12 (18) |
| Chronic lung disease | 17 (18) | 11 (17) |
| Chronic kidney disease | 4 (4) | 2 (3) |
| Congenital heart disease | 6 (6) | 3 (5) |
| Diabetes mellitus | 3 (3) | 1 (2) |
| Sickle cell disease | 1 (1) | 1 (2) |
| **Vaccine status** |  |  |
| Ineligible (<5 years of age) | 16 (17) | 12 (18) |
| Unknown | 1 (1) | 1 (2) |
| Eligible (5+ years of age) | 77 (82) | 53 (80) |
| 0 doses | 35 (37) | 28 (42) |
| 1 dose | 6 (6) | 6 (9) |
| 2 doses | 25 (27) | 15 (23) |
| 3 doses | 11 (12) | 4 (6) |
| **Not approved (N=28)\*\*** |  |  |
| Fully vaccinated*** | 13 (46) | N/A |
| Not in highest risk categories | 17 (61) |  |
| Too late in infection | 4 (14) |  |
| No treatment available | 1 (4) |  |
| Tixagevimab/cilgavimab recipient | 1 (4) |  |

\* The majority of those who chose “Other” race also chose Hispanic ethnicity
\*\* Patients may have more than one indication
\*\*\* Defined as $\geq 2$ doses of SARS-CoV-2 vaccination per Centers for Disease Control guidance

**Table 2.**
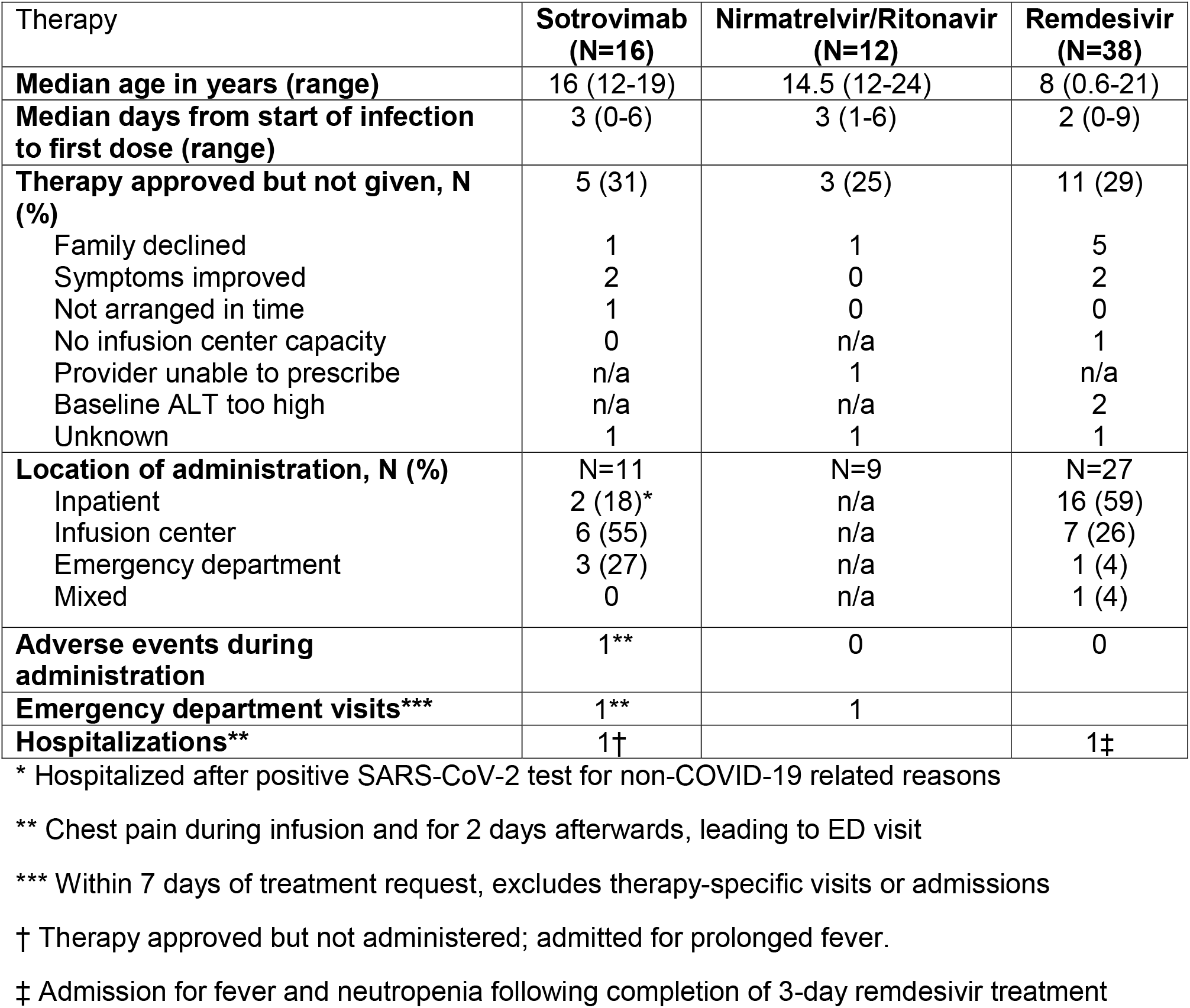
Administration characteristics and outcomes in patients approved for COVID-19 therapy.

| Therapy | Sotrovimab<br>(N=16) | Nirmatrelvir/Ritonavir<br>(N=12) | Remdesivir<br>(N=38) |
| --- | --- | --- | --- |
| **Median age in years (range)** | 16 (12-19) | 14.5 (12-24) | 8 (0.6-21) |
| **Median days from start of infection to first dose (range)** | 3 (0-6) | 3 (1-6) | 2 (0-9) |
| **Therapy approved but not given, N (%)** | 5 (31) | 3 (25) | 11 (29) |
| Family declined | 1 | 1 | 5 |
| Symptoms improved | 2 | 0 | 2 |
| Not arranged in time | 1 | 0 | 0 |
| No infusion center capacity | 0 | n/a | 1 |
| Provider unable to prescribe | n/a | 1 | n/a |
| Baseline ALT too high | n/a | n/a | 2 |
| Unknown | 1 | 1 | 1 |
| **Location of administration, N (%)** | N=11 | N=9 | N=27 |
| Inpatient | 2 (18)* | n/a | 16 (59) |
| Infusion center | 6 (55) | n/a | 7 (26) |
| Emergency department | 3 (27) | n/a | 1 (4) |
| Mixed | 0 | n/a | 1 (4) |
| **Adverse events during administration** | 1** | 0 | 0 |
| **Emergency department visits\*\*\*** | 1** | 1 |  |
| **Hospitalizations\*\*** | 1† |  | 1‡ |

\* Hospitalized after positive SARS-CoV-2 test for non-COVID-19 related reasons
\*\* Chest pain during infusion and for 2 days afterwards, leading to ED visit
\*\*\* Within 7 days of treatment request, excludes therapy-specific visits or admissions
† Therapy approved but not administered; admitted for prolonged fever.
‡ Admission for fever and neutropenia following completion of 3-day remdesivir treatment

## Discussion

This represents one of the first reports of early treatment of Covid-19 in high-risk children and adolescents infected with the Omicron variant of SARS-CoV-2. To the extent we can determine, all therapies were well tolerated and subsequent Covid-19 related ED visits or hospitalizations were uncommon.

While most children with Covid-19 do well without therapy, treatment of mild-to-moderate infection should be considered in those at highest risk of progression to severe disease. In fact, recent data demonstrates that hospitalization rates in children 0-4 increased significantly during Omicron data predominance, therefore information around early treatment in pediatric patients has become even more important [8]. However, despite FDA EUA for sotrovimab and nirmatrelvir/ritonavir in patients over 12 years and remdesivir in children < 12 years, safety and efficacy data for these agents in pediatric patients is extremely limited. The early treatment of remdesivir study included only 8 patients < 18 years of age [4].

Limitations of this study include its observational nature, small sample size at a single center, short follow up period and the possibility that not all adverse events and outcomes were reported in the medical record. As symptomatic patients with severe underlying conditions were more likely to have therapies requested and approved, true assessment of treatment impact on outcomes on a broader population is not possible. Though we saw no clear indicator of disparities in our approval process, we are doing further work to explore potential equity issues as have been seen in the distribution of Covid-19 therapies for adult populations nationwide [9].

## Data Availability

All data produced in the present study will be available upon reasonable request to the authors after peer review and publication of the final article.

## Contributors’ Statement Page

Dr.Vora conceptualized and designed the study, collected data and carried out the analysis, drafted the initial manuscript and reviewed and revised the manuscript.

Drs. Englund and Zerr conceptualized and designed the study and reviewed and revised the manuscript

Drs. Trehan, Waghmare and Kong reviewed and revised the manuscript.

Ms. Adler collected data and reviewed and revised the manuscript.

All authors approved the final manuscript as submitted and agree to be accountable for all aspects of the work.

